# A Common Data Model for the standardization of intensive care unit (ICU) medication features in artificial intelligence (AI) applications

**DOI:** 10.1101/2023.09.18.23295727

**Authors:** Andrea Sikora, Kelli Keats, David J. Murphy, John W. Devlin, Susan E. Smith, Brian Murray, Mitchell S. Buckley, Sandra Rowe, Lindsey Coppiano, Rishikesan Kamaleswaran, the MRC-ICU Investigator Team

## Abstract

**Objective:** Common Data Models provide a standard means of describing data for artificial intelligence (AI) applications, but this process has never been undertaken for medications used in the intensive care unit (ICU). We sought to develop a Common Data Model (CDM) for ICU medications to standardize the medication features needed to support future ICU AI efforts.

**Materials and Methods:** A 9-member, multi-professional team of ICU clinicians and AI experts conducted a 5-round modified Delphi process employing conference calls, web-based communication, and electronic surveys to define the most important medication features for AI efforts. Candidate ICU medication features were generated through group discussion and then independently scored by each team member based on relevance to ICU clinical decision-making and feasibility for collection and coding. A key consideration was to ensure the final ontology both distinguished unique medications and met Findable, Accessible, Interoperable, and Reusable (FAIR) guiding principles.

**Results:** Using a list of 889 ICU medications, the team initially generated 106 different medication features, and 71 were ranked as being core features for the CDM. Through this process, 106 medication features were assigned to two key feature domains: drug product-related (n=43) and clinical practice-related (n=63). Each feature included a standardized definition and suggested response values housed in the electronic data library. This CDM for ICU medications is available online.

**Discussion:** The CDM for ICU medications represents an important first step for the research community focused on exploring how AI can improve patient outcomes and will require ongoing engagement and refinement.

**Lay Summary:** Medication data pose a unique challenge for interpretation by artificial intelligence (AI) because of the alphanumerical combinations (e.g., ibuprofen 200mg every 4 hours) and because of the technical detail associated with drug prescriptions (e.g., ibuprofen 200mg and acetaminophen 325mg are both starting doses and round tablet sizes, so it would be incorrect for the machine to view 325mg as ‘more’ than 200mg). Because AI has great potential to improve the safety and efficacy of medication use, a common data model for ICU medications (ICURx) is proposed here to overcome these challenges and support AI efforts in medication analysis.

## Introduction

Artificial intelligence (AI) in critical care is a significant and rapidly growing field of investigation. [1–8] However, for AI to improve the outcomes of critically ill patients, precise feature selection and standardized, machine-readable ontologies are necessary to create robust, well-validated models. [9 10] A significant challenge facing the AI community is creating standardized ontologies for the different components of datasets (e.g., laboratory values, vital signs, clinical interventions, etc.). For example, when data are pulled from data warehouses regarding serum bicarbonate values, these may read as ‘serum bicarbonate,’ ‘bicarbonate,’ ‘CO_2_,’ ‘HCO_3_,’ or any number of institution-specific monikers). Without standardization, an AI algorithm may interpret each of these values differently, which can be prohibitive for validating algorithms in external datasets. Standardization to ensure each laboratory test result (or clinical intervention) will always be referred to by the same name (and ideally the same unit of value or exposure) requires a common data model (CDM) to facilitate both comparisons among datasets and adaptation and improvement of algorithms and models among different investigator teams.

A CDM refers to any data model generally consisting of an ontology and associated metadata that allows for standardized data and information exchange among different applications and data sources. CDMs facilitate reproducibility and generalizability across datasets, and without these efforts, external validity of a significant portion of research efforts in AI are jeopardized. [11] The importance of these CDM development efforts have been internationally recognized with the publication of the FAIR Guiding Principles, which are intended to steward scientific data to be Findable, Accessible, Interoperable, and Reusable. [12]

Complex medication regimens are frequently used in the intensive care unit (ICU) when treating patients; medications play an important role in determining patient outcomes. [13 14] While ICU medication characterization is a necessary component of AI models, to date, only simplistic, non-ICU based common data models (limited to medication name, dose, and route) have been developed. [9 15 16] Underlying patient features affecting drug response and clinical outcomes denoting medication titration, efficacy and/or safety have not been considered. The current lack of clinically relevant medication information in contemporary ICU AI models may result in an AI algorithm viewing different medication orders the same way despite the indication for use being drastically different and the pharmacokinetic/pharmacodynamic effects being distinctive. For example, the same heparin product may be given for both routine prophylaxis of venous thromboembolism or the management of a life-threatening pulmonary embolism. Thus, without a rigorously enhanced CDM for ICU medications the ability to use AI to support medication therapy optimization remains limited.

We sought to develop a CDM for ICU medications to standardize the medication features needed to support future ICU AI applications.

## Materials and Methods

### Design

A consensus process based on a modified Delphi approach was initiated in April 2022. This method has been successfully applied in healthcare research efforts including common data element development, prescribing guideline generation, and medication outcome prioritization. [17–21] Choosing this approach allowed the team to address temporal and cost constraints while ensuring each individual subject matter expert had an equal voice. [22]

### Participants

The 9-member expert panel was invited to participate in a 5-round modified Delphi process, including one computer scientist, one intensivist, and seven critical care pharmacists. All eight clinicians represented geographically diverse locations and held critical care board-certification in their respective professions; each were therefore deemed content experts in the domain of critical care pharmacotherapy and able to cast votes regarding the clinical significance all CDM attributes. The computer scientist contributed to framing the process and providing important insights for CDMs and deferred on clinical considerations. The University of Georgia Institutional Review Board (IRB) deemed this project to be exempt from IRB review (PROJECT00006204). Voluntary involvement in the Delphi process implied participant consent. ***Framework***

Participants were briefed on the primary goal of the expert panel: to develop a CDM for ICU medications for use in AI applications. A framework consisting of concept, purpose, and ideal characteristics for this CDM was supplied to the panel to facilitate a consistent development process (**see Table 1**). *A priori,* it was defined each CDM would be comprised of a list of medication features and standardized definitions and coded suggestions. In machine learning, a feature can be thought of as an individual, measurable property or characteristic of a particular entity. For any given medication, an individual feature could include dose, route, the risk for drug-drug interactions, etc. [23] The group was asked to consider various situations focused on how ICU clinicians delineate unique features among different medications used in the ICU (see **Table 2**).

**Table 1.** Framework for development of the Common Data Model for ICU Medications.

|  |
| --- |
| **Concept** |
| ? For use in artificial intelligence and other data science applications investigating critical illness |
| **Purpose** |
| ? Provide an intensive care unit medication ontology that meets the Findable, Accessible, Interoperable, and Reusable (FAIR) Guiding Principles |
| **Characteristics** |
| ? Ability to distinguish unique medication identities and profiles |
| ? Captures relevant medication features used in clinical decision making |
| ? Feasible to incorporate into more definitive database |

**Table 2.**
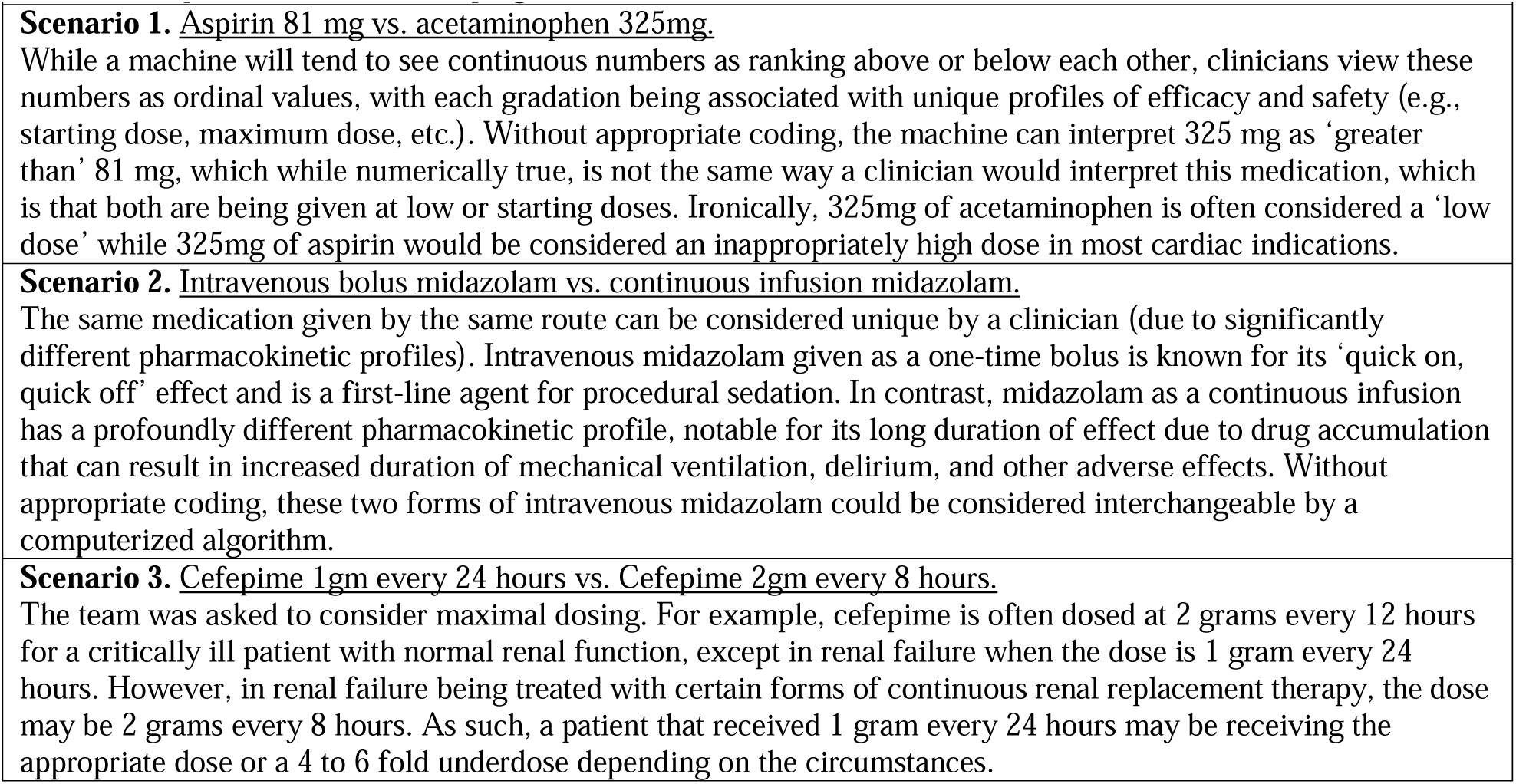
Proposed cases for developing machine-readable ICU medication features.

### Study sample

A representative ICU medication list was derived from the unified electronic health record (EHR) from a random sample of 1,000 ICU patients at the University of North Carolina Health System between October 2015 - October 2020. Inclusion criteria for patients consisted of age ≥18 years old on their index ICU admission who had been admitted to the burn, cardiac, cardiothoracic surgery, medical, mixed, neurosciences, or surgical ICU. Exclusion criteria included patients with active comfort care orders.

Given the Federal Food and Drug Administration (FDA) has approved over 20,000 *medication products* and that each individual medication product can be ordered in a myriad of unique ways, a nearly infinite number of *medication orders* exists. For example, the medication product of ‘aspirin 325mg tablet’ may be ordered as once daily, three times daily, three times daily as needed for headache, etc. For the feasibility of this initial process, the team focused on two key areas: first, medication products (e.g., aspirin 325mg tablet) and second, medication products mostly likely to be used in the ICU as defined by a previously validated metric (the medication regimen intensity-intensive care unit (MRC-ICU) Score). [13 14 24-31] Because these medications in the MRC-ICU have been previously identified as common to ICU care and having unique characteristics that make their use associated with increased ICU patient care complexity and a requirement for expert oversight, they were deemed an appropriate initial list of representative medications for CDM development. [13 14 24-31]

### Modified Delphi Process

The modified Delphi process is depicted in **Figure 1**. Prior to the virtual meeting for Round 1, a description of the objective and an initial list of 50 potential medication features were electronically sent to each participate to provide a starting point for independent brainstorming. In Round 1, the goal was to compile and define an initial series of machine-readable features (or common data elements) that characterize medications and influence ICU clinical decision-making. Each member of the panel was provided the opportunity to independently review an initial list of features, provide feedback, and suggest additional features. All Round 1 feedback from the expert panel was evaluated and incorporated into a revised library for second round deliberation.

**Figure 1.**
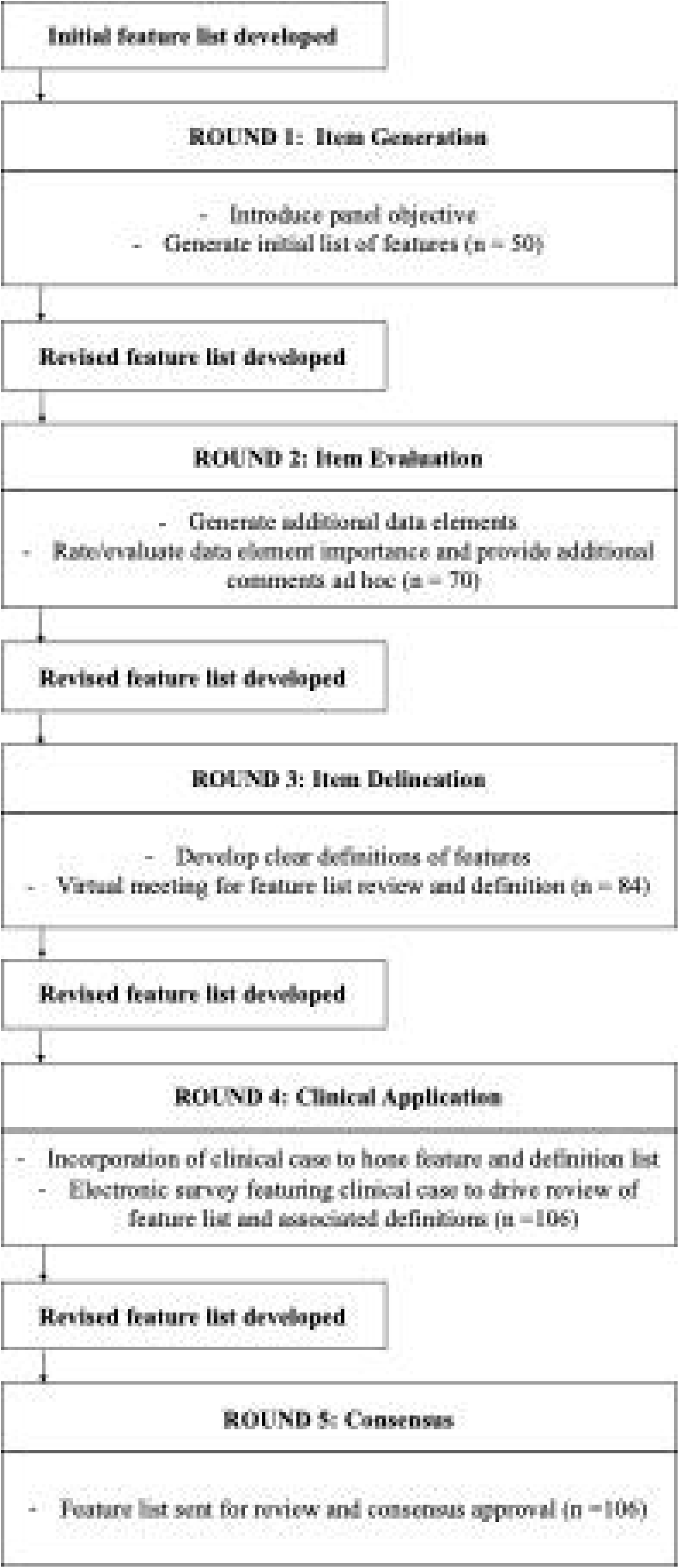
Modified Delphi process for development of the Common Data Model for ICU Medications.

In Round 2, the goal was additional item generation. An electronic survey was administered regarding feature importance (see **Appendix Table 1**). A survey, intended to be completed independently and anonymously, was provided to each panel member to vote on the relative importance of each feature in the library (see **Appendix Table 1**). Comments about new features and feedback on existing features was also encouraged. The results of this survey were incorporated into a new list of common data elements and distributed to each member.

In Round 3 of the Delphi process, the expert panel met to review and discuss the results of Round 1 and 2, including each of the new features added. The goal of this round was to identify broad themes of the common data elements generated and prioritize which elements should be included in the first CDM version.

Following this in-group discussion, Round 4 consisted review and comment regarding an updated list of medication features. In this round, a patient case was presented that included a medication regimen. The prompts asked the panel to consider features “that would be most relevant and most important to clinical decision-making associated with comprehensive medication management of that patient” and vote for inclusion into three categories: (1) a <u>core list</u> of features for use in ICU AI algorithms not specifically focused on medical use; (2) a <u>core list</u> of features for use in ICU AI algorithms targeted to medication use optimization; and (3) an <u>expanded list</u> of features for use in ICU AI algorithms targeted to medication use optimization. While feasibility and parsimony were considered, they were deemed secondary priorities given the general construct of AI is to maximize the use of available data and the ultimate goal of this library was for it to be able to be readily linked to any investigator’s dataset. Inclusion in the first category was defined as having greater than or equal to 4 votes (see **Appendix Table 2**).

In Round 5, a final list of features and their definitions was sent out for approval that the panel believed adequately represented key features of medications used in the ICU. Approval was indicated by anonymous voting. The list of Core Features is provided in **Table 3**. The complete list of features and their associated definitions is provided in **Appendix Table 3**. The data underlying this article are available in GitHub, under ICURx at: https://www.icurxforai.com/.

**Table 3.** List of medication features.

| **Table 3. List of medication features** |  |
| --- | --- |
| **Drug Product Features** |  |
| • Drug Product (Core)<br>• Drug Name (Core)<br>• Therapeutic Category (Core)<br>• Pharmacologic Class (Core)<br>• Formulary Key Drug Type (Expanded)<br>• Formulation Strength (Core)<br>• Formulation Strength Conversion (Expanded)<br>• Converted Drug Name (Core) | • Converted Drug Strength (Core)<br>• Strength Units (Core)<br>• Route (Core)<br>• Intravenous volume (Expanded)<br>• Concentration (Expanded)<br>• Intravenous, sub-category (Core)<br>• Oral, sub-category (Core) |
| **Clinical Features** |  |
| **Dosing Information** | **Laboratory Monitoring** |
| • Fixed Route (expanded)<br>• Continuous Infusion Starting Rate (Core)<br>• Continuous Infusion Maximum Rate (Core)<br>• Units for continuous infusion (Core)<br>• Weight-based Dosing (Core)<br>• Maximum Daily Dose Available (Core)<br>• Maximum Daily Dose Value (Core)<br>• Maximum Daily Strength Units (Core)<br>• Maximum daily dose for Creatine Clearance < 10 mL/min or dialysis (Core)<br>• Maximum daily dose for Creatine Clearance = 10-29 mL/min (Core)<br>• Maximum daily dose for Creatine Clearance = 30-49 mL/min (Core)<br>• Maximum daily dose for Creatine Clearance < 10 mL/min or dialysis and critically ill (Expanded)<br>• Maximum daily dose for Creatine Clearance = 10-29 mL/min and critically ill (Expanded)<br>• Maximum daily dose for Creatine Clearance = 30-49 mL/min and critically ill (Expanded)<br>• Maximum daily dose for Creatine Clearance < 10 mL/min or dialysis and has cystic fibrosis (Expanded)<br>• Maximum daily dose for Creatine Clearance = 10-29 mL/min and has cystic fibrosis (Expanded)<br>• Maximum daily dose for Creatine Clearance = 30-49 mL/min and has cystic fibrosis (Expanded)<br>• Renal Dose Adjustment (Core)<br>• Hepatic Dose Adjustment (Core)<br>• Contraindicated in Renal Failure (Core)<br>• Contraindicated in Hepatic Failure (Core)<br>• Role in prophylaxis (Expanded)<br>• Maximum prophylactic dose (Expanded) | • Sodium (Core)<br>• Potassium (Core)<br>• Serum Creatinine (Core)<br>• Glucose (Core)<br>• Aspartate Aminotransferase (Core)<br>• Alanine Transaminase (Core)<br>• Magnesium (Core)<br>• Creatinine Kinase (Core)<br>• Albumin (Core)<br>• Triglycerides, lipid panel (Core)<br>• Activated partial thromboplastin time (Core)<br>• International normalized ratio (INR) (Core)<br>• Anti-Xa (Core)<br>• Hemoglobin (Core)<br>• Hematocrit (Core)<br>• Platelets (Core)<br>• Absolute neutrophil count (Core)<br>• White blood cell count (Core)<br>• Microbiology results (cultures, sensitivity, rapid results) (Core)<br>• Phosphorous (Expanded)<br>• Serum osmolality (Expanded)<br>• Ammonia (Expanded) |
| **Vital Sign and other ICU Monitoring** | **Clinical Decision-Making** |
| • Mean arterial pressure (Core)<br>• Systolic blood pressure (Core)<br>• Diastolic blood pressure (Core)<br>• Heart Rate (Core)<br>• Respiratory rate (Core)<br>• Temperature (Core) | • MRC-ICU Presence (Core)<br>• MRC-ICU Weight (Core)<br>• Route Escalation (Core)<br>• Patient Acuity (Core)<br>• Critical Illness Based Dosing (Core)<br>• ISMP High Alert Status (Core) |
| • Creatinine Clearance (Core)<br>• Urine output (Core)<br>• Dialysis (Core)<br>• Patient Weight (Core)<br>• QTc Interval (Core)<br>• Bowel Movements (Core)<br>• Train of Four (Core)<br>• Pain Scale (Core)<br>• Sedation scale (Core)<br>• Delirium (CAM-ICU) (Core) | • ISMP Confused Drug Names (Core)<br>• Therapeutic Drug Monitoring (Core)<br>• Parameter Based Dosing (Core)<br>• CYP3A4, 2D6, 1A2, 2C9, 2C19, P-glycoprotein Interaction (Core)<br>• Drug Indicates Critical Illness (Core)<br>• Adverse events (Expanded)<br>• Realm of the unusual (Expanded)<br>• Beers Criteria (Expanded)<br>• Beers Criteria Strong Recommendation (Expanded)<br>• Pharmacogenomics (Expanded)<br>• Broad vs. Narrow Spectrum (Expanded)<br>• Vancomycin-Resistant *Enterococcus spp.* (Expanded)<br>• Methicillin-Resistant *Staphylococcus Aureus* (Expanded)<br>• *Pseudomonas aeruginosa* (Expanded)<br>• Extended Spectrum Beta Lactamase (Expanded)<br>• Treatment Failure (Expanded) |

## Results

### Study sample

From the random sample of 1,000 patients, 9 patients were excluded due to being not the index ICU admission. From here, the derived medication list comprised of 30,550 discrete medication order entries for 991 ICU patients. [24] When a filter for the generic drug name, dose, and administration route was applied, a total of 1,868 unique medication products were identified. When only those medications incorporated in the MRC-ICU Scoring Tool were considered, a total of 889 discrete medication products remained for review and coding by the panel.

### Modified Delphi Process

As a result of Round 1 and the subsequent electronic discussion, the medication feature list expanded from 50 to 70. After the end of the virtual meeting for Round 3, a total of 84 features (along with definitions) was generated. Upon completion of the case in Round 4, features were expanded to a total of 106 (see **Appendix Table 2**). Following voting, a total of 71 core features were identified (see **Table 3**). The final CDM approved by the panel can be found in **Appendix Table 3**).

Our process revealed three distinct feature domains: 1) <u>Drug product features</u>: nature of the product used including drug name, drug dose, formulation and route of administration; 2) <u>Clinical features</u>: medication characteristics driving ICU clinical-decision making regarding drug product use (e.g., the risk for a known adverse drug event in the setting of critical illness); and, 3: <u>Order features</u>: specific way in which the drug product was ordered for the patient (i.e., the ‘sig’) (e.g., acetaminophen 325mg oral tablets could be ordered on a scheduled basis as two tablets every 6 hours or on a ‘prn” basis as two tablets every 4 hours as needed for a pain score ≥ 4/10). Given the construction of this medication product list which did not include medication orders and their associated features, only the Drug Product Feature and Clinical Feature domains were further developed; however, this area was noted to be an important area of future research as clinicians may view medications differently when viewed ‘in a silo’’ vs. in context of the entire medication profile or the specific order. Within the clinical feature domain, four medication product-related feature categories were identified: dosing, laboratory monitoring, clinical monitoring (including vital signs), and clinical-decision making.

### Common Data Model

The CDM for ICU medications is housed electronically (icurxforai.com). It consists of 94,234 coded elements for 889 medications and 106 medication features. A data dictionary defines and expands upon the various features and provides standardized coding.

## Discussion

This paper describes the development of the first CDM focused on medications used in the ICU and includes a medication library of 889 ICU medications described by 106 individual features. Our efforts represent the first CDM medication development to prioritize clinical features through an expert, modified Delphi process and represents an important next step for AI in the ICU, which has significant potential to improve patient outcomes via both prediction of ICU complications as well as via clinical decision support systems (CDSS).

Medications represent a particularly important domain for critical care predictive algorithms and CDSS because they serve as both treatments to life-threatening illness but also independent risk factors for life-threatening complications due to adverse drug events (ADEs). [13 32 33] Medications in the ICU setting are uniquely challenging to optimally manage because many are high-risk (i.e., beneficial effects are also associated with important potential safety concerns) and often have a narrow-therapeutic index (i.e., a narrow difference in the medication dose resulting in benefit vs. side effects). The fact that medications in the ICU setting can be used in a nearly infinite number of combinations often results in the creation of complex, multi-drug medication regimens necessitating constant optimization by critical care clinician experts.

Key ICU medication clinical decision-making components include drugs with synergistic mechanisms, overlapping side effect profiles resulting ADE risks that are additive, and the fluctuating nature of critical illness resulting in frequent pharmacokinetic and pharmacodynamic alterations. [16] While ICU clinicians routinely apply their nuanced knowledge of ICU medication features to efficiently manage the benefits and harms of medication therapy, this process has had no equivalent in the AI domain given its unstructured and complex nature that precludes the creation of robust machine-readable libraries. [16]

Current medication CDM efforts including the RxNorm by the National Library of Medicine [34] and the Observational Medical Outcomes Partnership (OMOP) [35] CDM have prioritized reproducibility when organizing medication products and have not yet considered many of the more clinical aspects of medication prescribing and outcomes. The primary goal of RxNorm is to normalize drug products using identifying codes (e.g., “Albuterol 0.417 MG/ML Inhalation Solution” coded 351136). [36] Realizing the FDA has approved more than 20,000 drug products the efforts of RxNorm are commendable; however, the provision of relevant features needed for clinical-decision making represents a key next step in this infrastructure that will further support CDSS development.

Acknowledging the complexity of ICU comprehensive medication management optimization, our effort sought to capture the nuances of clinical-decision making and incorporate it into a CDM through a multiple-step, consensus-based process involving an interprofessional group of experienced ICU clinicians and an AI methodologist. Our Delphi process allowed for nuanced discussion of the ICU medication clinical decision-making process not captured in the package insert or other standardized drug references. A notable strength of this process included grounding CDM development discussions in patient case scenarios. This process resulted in all medication features coded and defined as a CDM that extends beyond drug product mappings of present day CDMs to the realm of clinical decision-making for use in ICU AI applications.

Incorporation of ICU medications into AI algorithms are in the early stages of development. A variety of algorithms have been previously developed that included indicators of a broad class of medications, such as use of inotropes and vasopressors, without adjusting for the individual’s overall regimen.[40–47] While such broad strokes were often used due to limitations of the technology, it is salient to realize that large amounts of relevant detail were ignored, given the unique pharmacokinetic and pharmacodynamic profile of each drug and that they can interact with other drugs in ways that go unaccounted for with that approach. Unsupervised machine learning using Restricted Boltzmann Machine has been applied to the medication administration records of ICU patients revealing unique pharmacophenotypes associated with patient outcomes. The first iteration used just generic drug name in addition to basic patient demographics and ICU outcomes, excluding all other potential drug features, but still showed the presence of pharmacophenotypes.[24] The second employed a significantly expanded list of drug features in line with this proposed CDM, showing again the ability of AI to incorporate vast amounts of data into its predictive algorithms.[40] Understanding how to translate these pharmacophenotypes into clinically meaningful subgroups or interventions for a bedside clinician remains an ongoing area of investigation. Medication data have also been incorporated into supervised learning for prediction of prolonged duration of mechanical ventilation and mortality; however, neither of these analyses included the entire ICU medication regimen and focused just on generic name.[28,41,43] Similarly, a large analysis of factors associated with hemodynamic compromise did incorporate some medication data, although it was not the complete regimen.[42] A common theme is the improvement of modeling performance with the inclusion of medication data (particularly drug name or class), though little has been done incorporating comprehensive medication data for the patient’s regimen (including both all medications they received and relevant information on those medications) to date.

Despite the strengths of our process, several potential limitations exist. First, our process did not include all available medications used in the ICU, which will be an important area of future development. Second, the feature profile for each medication feature may not always complete (e.g., possible non-cytochrome P450 drug-drug interactions that could influence medication response) and while development of such features was considered out of scope for this initial process, additions are likely warranted. Finally, although great care was taken to ensure proper coding, some of the CDM features are based on the expert opinion of our panel, which may differ from the viewpoints of ICU clinicians who were not involved.

We consider our proposed CDM for ICU Medications CDM to be a living and evolving database: iterative additions and revisions will be required as new medications are discovered and the ICU drug literature expands. Several next steps are proposed. First, given the present CDM does not include patient-level, order-specific or case-specific temporal features, future research methodology focused on how the nuances of medication orders is warranted. This research should focus on how combinations of medications may prompt increased scrutiny or medication order modification by ICU clinicians. Indeed, pattern-recognition based clinical reasoning models have been proposed in the pedagogy of medical education, and, already, the concept of AI-assisted pattern recognition for diagnosis is being investigated. [37 38] Just as experience with a constellation of patient signs and symptoms can lead to rapid recognition of a new ICU clinical diagnosis, medication use also has certain patterns. [38 39] When an ICU clinician reviews the medication profile, each individual medication order is evaluated in a broader clinical context with the other medication orders. For example, a patient with one active vasopressor order likely registers very differently different to the clinician than a patient requiring three vasopressors. Thus, in the context of comprehensive medication management, clinicians become increasingly attuned to ‘the realm of the unusual’ for any medication regimen. Encoding medical expertise with some degree of level of evidence or strength of evidence, as is seen in clinical guidelines, could serve as a useful future addition to add further nuance to AI algorithms. Given that different use cases (e.g., workload allocation, ADE prediction) likely require different datapoints, it is likely that ‘one size does not fit all’ and different iterations of features may be needed, although these iterations require separate methodologies than used here.

## Conclusion

A modified Delphi process was applied to develop an initial standardized ontology of ICU medications for future use in AI research. This work represents a first step towards developing a CDM, and future research may focus on evaluation on validation and relevance of these features for making clinically relevant predictions in the ICU.

## Appendix I

**Table 1.** Respondent Votes for Round 2 Survey Questions.

| What is the level of priority for inclusion of this medication feature into a Common Data Model for ICU Medications? |  |  |  |  |
| --- | --- | --- | --- | --- |
|  | High Priority | Low Priority | Do not Include | Comment |
| Drug-Strength-Formulation (e.g., acyclovir 200mg capsule) | 4 | 4 | 0 | 0 |
| Drug Name (e.g., acyclovir) | 8 | 0 | 0 | 0 |
| Pharmaceutical class (e.g., antiviral agent) | 7 | 1 | 0 | 0 |
| Pharmaceutical subclass (e.g., DNA polymerase inhibitor) | 5 | 3 | 0 | 0 |
| Dose (e.g., 200) | 6 | 2 | 0 | 0 |
| Dose units (e.g., mg) | 5 | 3 | 0 | 0 |
| Volume, if intravenous (e.g., 100) | 3 | 4 | 1 | 0 |
| Concentration (e.g., 1mg/1mL) | 4 | 3 | 1 | 0 |
| Route (e.g., oral, intravenous) | 8 | 0 | 0 | 0 |
| IV subcategory (e.g., bolus vs. continuous infusion) | 7 | 1 | 0 | 0 |
| Bolus sub-category (e.g., IV push vs IV piggyback) | 4 | 3 | 1 | 0 |
| Oral sub-category (e.g., capsule, suspension) | 2 | 5 | 0 | 1 |
| Included in MRC-ICU | 6 | 1 | 0 | 1 |
| MRC-ICU Weight (i.e., 1,2,3) | 6 | 1 | 0 | 1 |
| Fixed route (i.e., only comes as oral agent) | 3 | 4 | 1 | 0 |
| Route escalation (e.g., IV instead of PO for same agent) | 6 | 2 | 0 | 0 |
| Therapeutic drug monitoring availability | 8 | 0 | 0 | 0 |
| Relative importance of TDM in the ICU | 7 | 1 | 0 | 0 |
| Cytochrome P450 Enzyme | 6 | 1 | 1 | 0 |
| Pharmacogenomics | 3 | 4 | 1 | 0 |
| Dosing alterations are present due to critical illness | 7 | 0 | 0 | 1 |
| Drug is indicative critical illness | 7 | 0 | 0 | 1 |
| Weight-based dosing is used | 7 | 1 | 0 | 0 |
| Maximum daily dose | 5 | 2 | 1 | 0 |
| Maximum daily dose adjusted by levels of creatinine clearance | 5 | 2 | 1 | 0 |
| Renal dose adjustment | 6 | 2 | 0 | 0 |
| Hepatic dose adjustment | 6 | 2 | 0 | 0 |
| Contraindication in renal failure | 6 | 2 | 0 | 0 |
| Contraindication in hepatic failure | 6 | 2 | 0 | 0 |
| Used for prophylaxis of disease | 4 | 4 | 0 | 0 |
| Institute for Safe Medication Practices (ISMP) High Alert Drug List<sup>1</sup> | 3 | 4 | 1 | 0 |
| ISMP Confused Drug List<sup>2</sup> | 1 | 6 | 1 | 0 |
| Beers Criteria<sup>3</sup> | 1 | 6 | 1 | 0 |
| Continuous Infusion Starting Rate | 5 | 2 | 1 | 0 |
| Continuous Infusion Maximum Rate | 5 | 2 | 1 | 0 |
| Continuous Infusion units | 5 | 3 | 0 | 0 |
| What is the level of priority for inclusion of this medication monitoring feature into a Common Data Model for ICU Medications? |  |  |  |  |
| Creatinine Clearance | 7 | 1 | 0 | 0 |
| Serum creatinine | 6 | 2 | 0 | 0 |
| Urine output | 6 | 2 | 0 | 0 |
| Dialysis | 8 | 0 | 0 | 0 |
| Sodium | 5 | 3 | 0 | 0 |
| Potassium | 4 | 4 | 0 | 0 |
| Chloride | 3 | 5 | 0 | 0 |
| Bicarbonate | 5 | 3 | 0 | 0 |
| Blood urea nitrogen | 4 | 4 | 0 | 0 |
| Glucose | 6 | 2 | 0 | 0 |
| Magnesium | 6 | 2 | 0 | 0 |
| Phosphate | 5 | 3 | 0 | 0 |
| Liver function tests | 6 | 2 | 0 | 0 |
| Creatinine kinase | 5 | 3 | 0 | 0 |
| Cultures/Sensitivities | 8 | 0 | 0 | 0 |
| activated partial thromboplastin time (APTT) | 7 | 1 | 0 | 0 |
| Anti-Xa levels | 8 | 0 | 0 | 0 |
| Prothrombin time/international normalized ratio | 7 | 1 | 0 | 0 |
| Hemoglobin | 6 | 2 | 0 | 0 |
| Hematocrit | 4 | 4 | 0 | 0 |
| Platelets | 6 | 2 | 0 | 0 |
| White blood cell count | 6 | 2 | 0 | 0 |
| Patient weight | 7 | 1 | 0 | 0 |
| QTc Interval | 7 | 0 | 0 | 0 |
| Serum Osmolality | 4 | 4 | 0 | 0 |
| Ammonia | 4 | 4 | 0 | 0 |
| Absolute neutrophil count | 5 | 3 | 0 | 0 |
| Train of Four | 6 | 1 | 0 | 0 |
| Critical Care Pain Observation Tool or other Pain Score | 8 | 0 | 0 | 0 |
| Richmond Agitation and Sedation Scale (RASS) or other agitation/sedation score | 8 | 0 | 0 | 0 |
| Confusion Assessment Method for the Intensive Care Unit (CAM-ICU) | 7 | 1 | 0 | 0 |
| Mean arterial pressure | 8 | 0 | 0 | 0 |
| Heart Rate | 8 | 0 | 0 | 0 |
| Systolic blood pressure | 6 | 1 | 0 | 0 |
| Respiratory rate | 7 | 1 | 0 | 0 |
| 1. https://www.ismp.org/sites/default/files/attachments/2018-08/highAlert2018-Acute-Final.pdf. Accessed 11/17/22.<br>2. https://www.ismp.org/recommendations/confused-drug-names-list. Accessed 11/17/22.<br>3. By the 2019 American Geriatrics Society Beers Criteria® Update Expert Panel. American Geriatrics Society 2019 Updated AGS Beers Criteria® for Potentially Inappropriate Medication Use in Older Adults. J Am Geriatr Soc. 2019 Apr;67(4):674-694. |  |  |  |  |

**Table 2.**
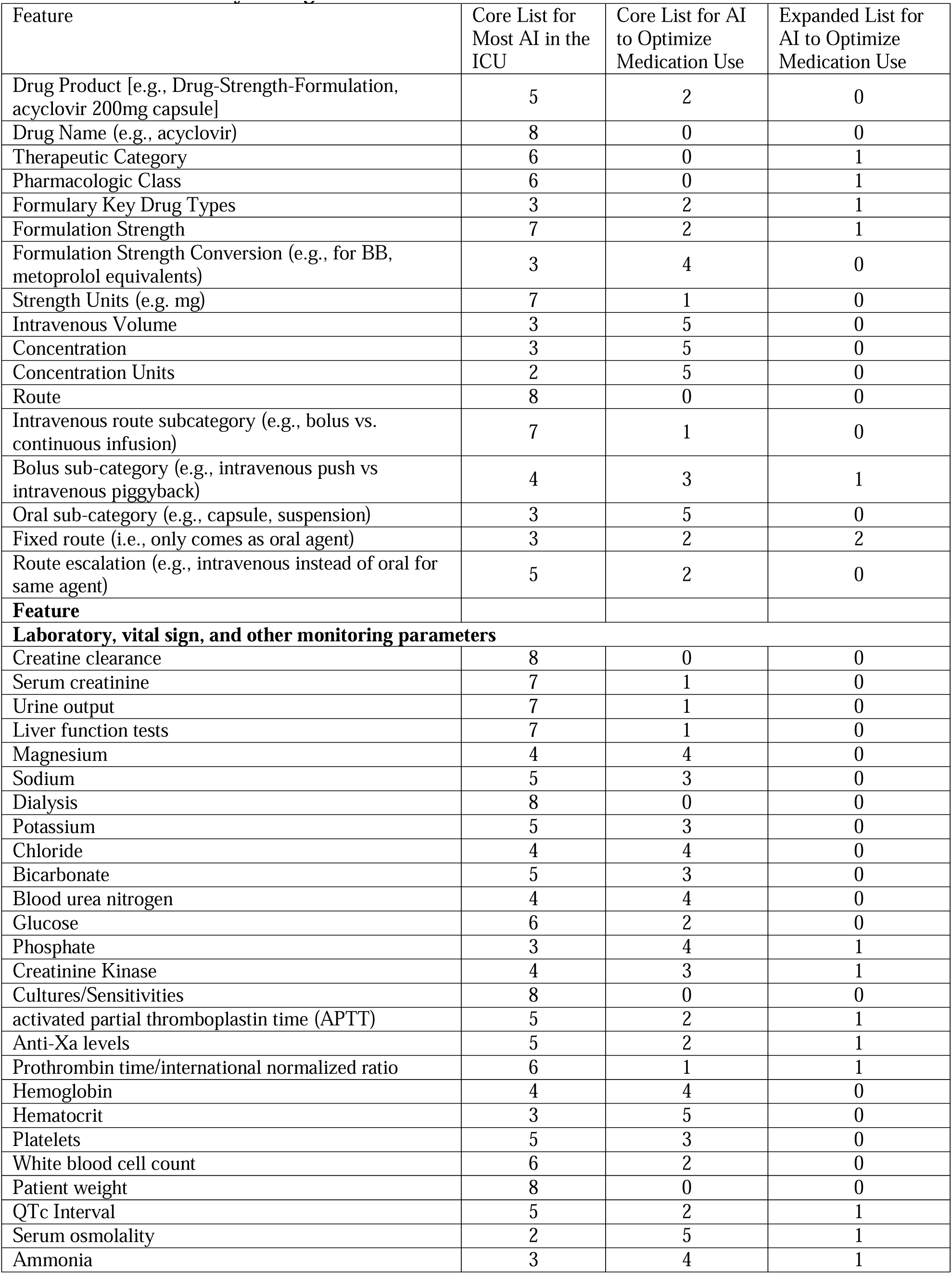
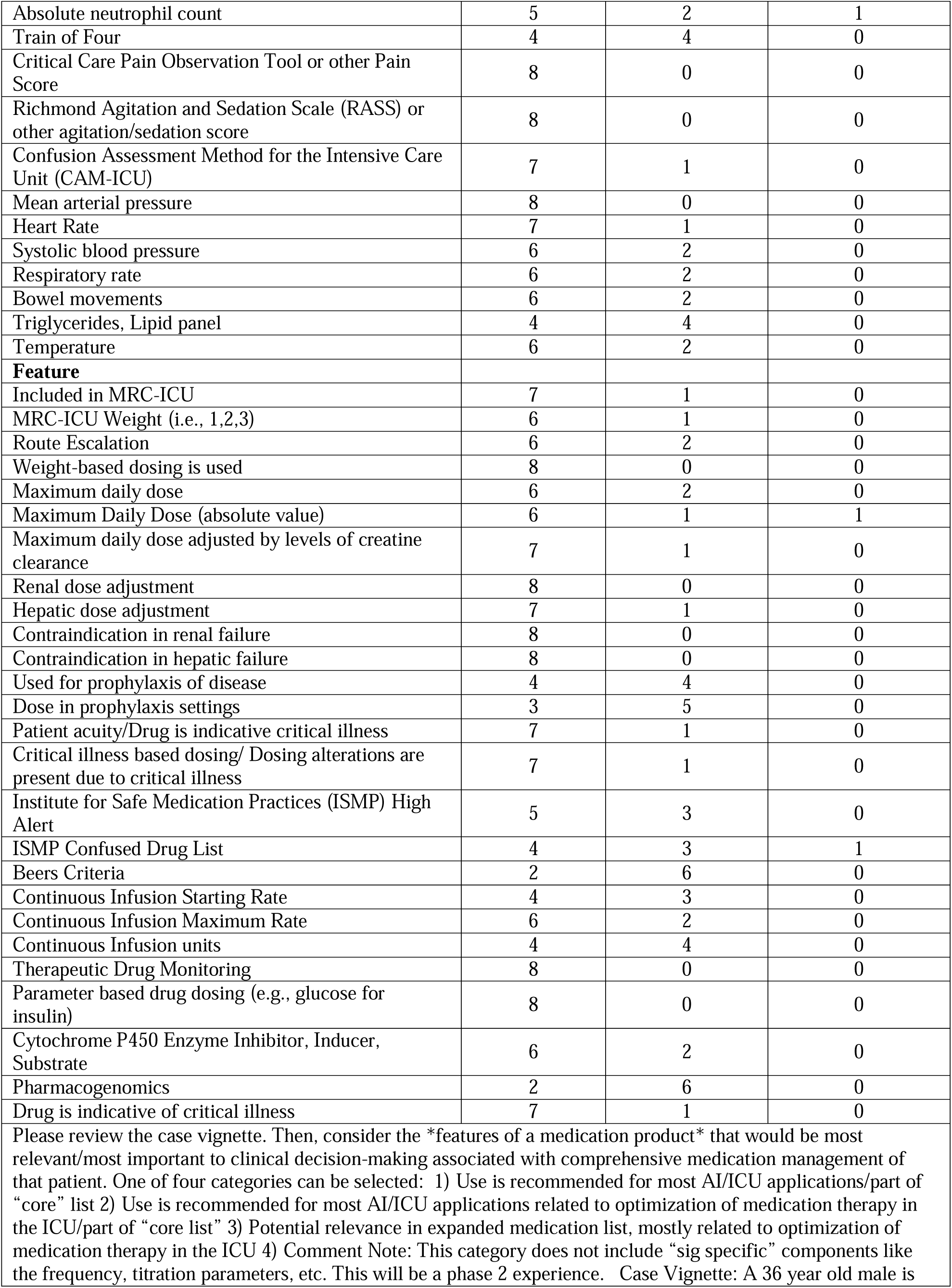
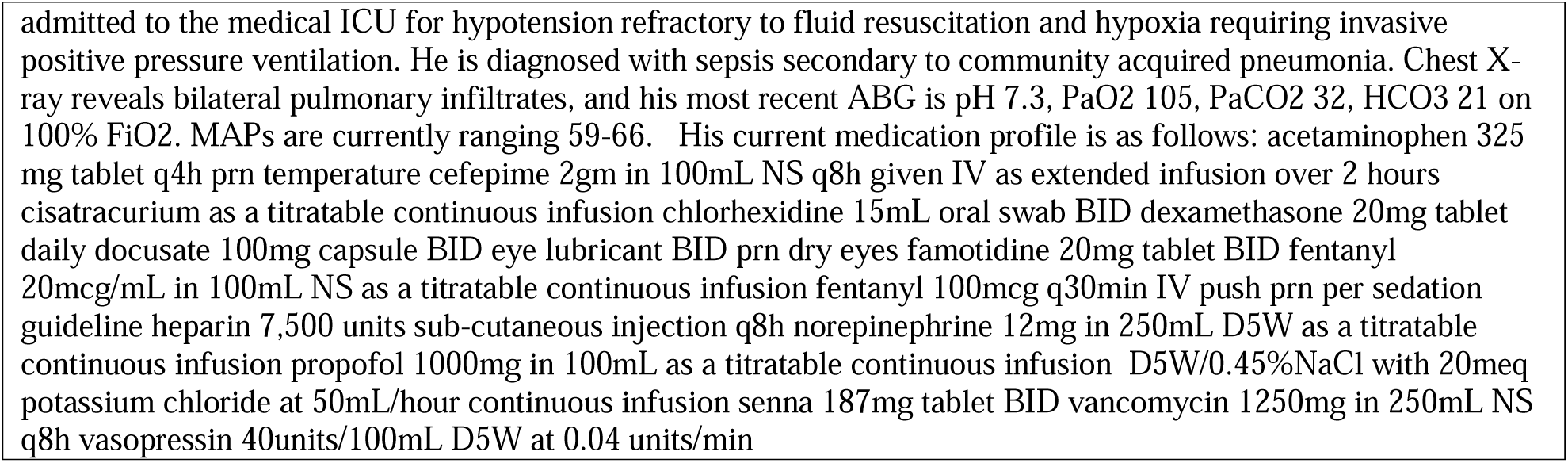
Round 4 Survey Voting Results.

**Table 3.**
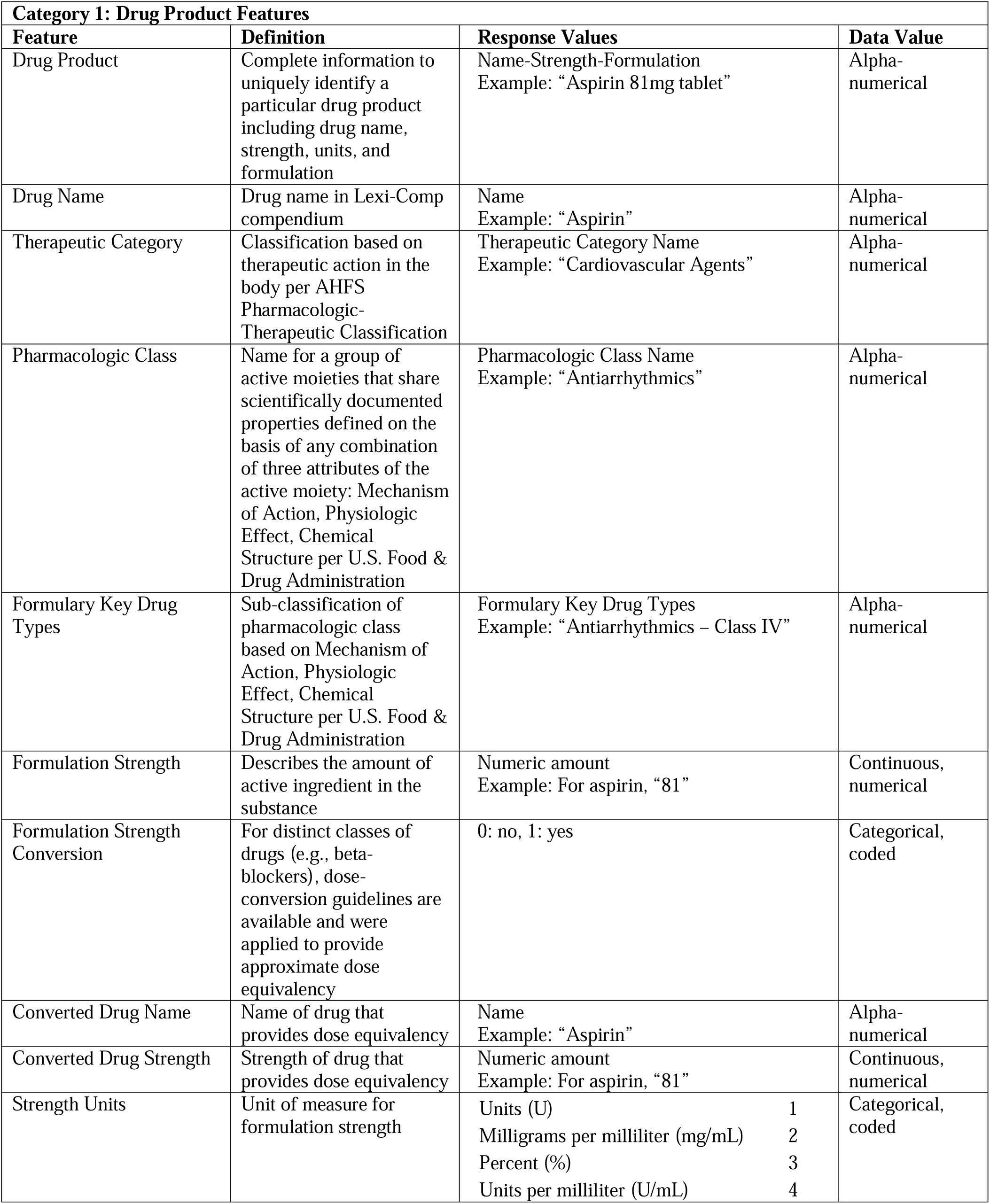
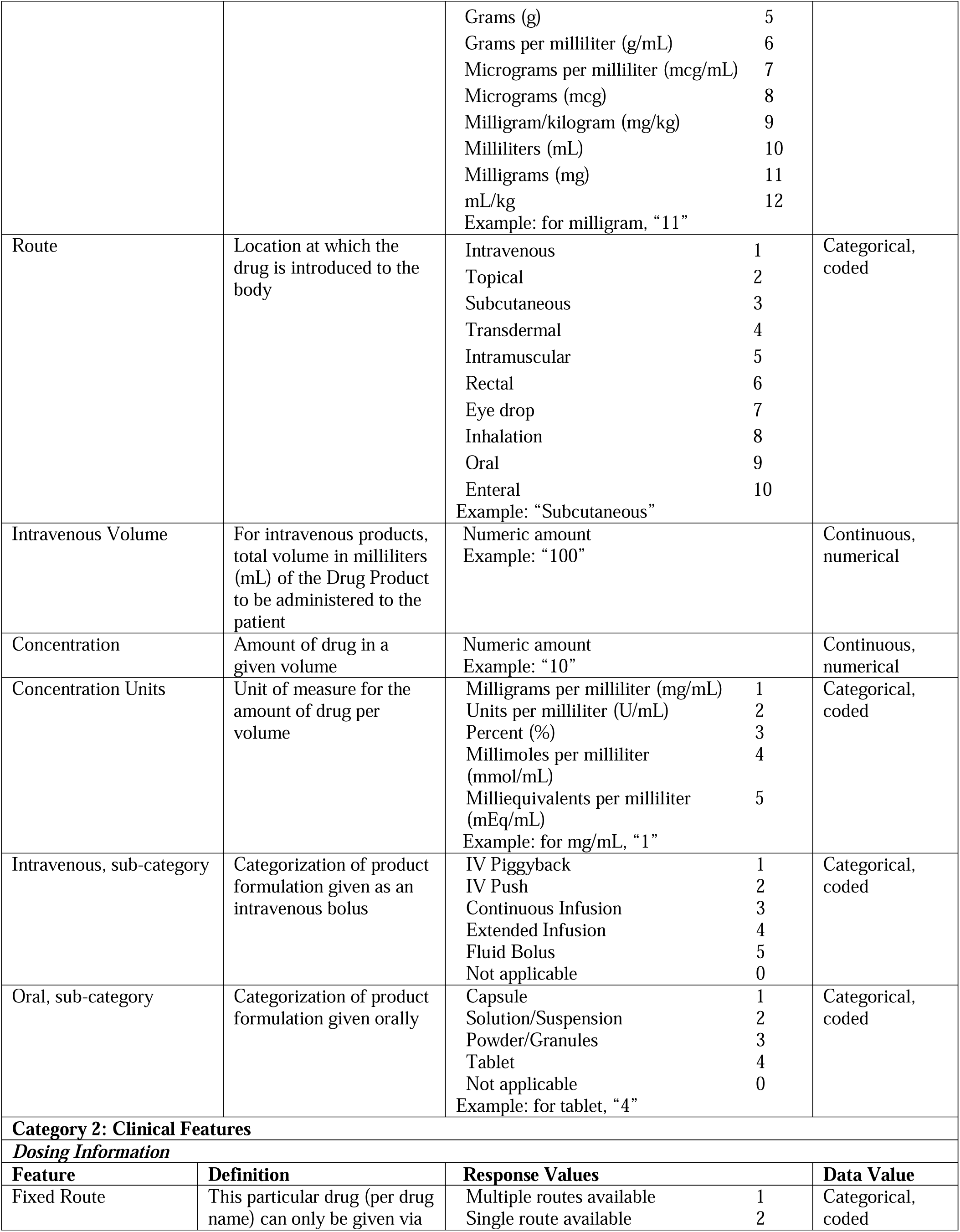
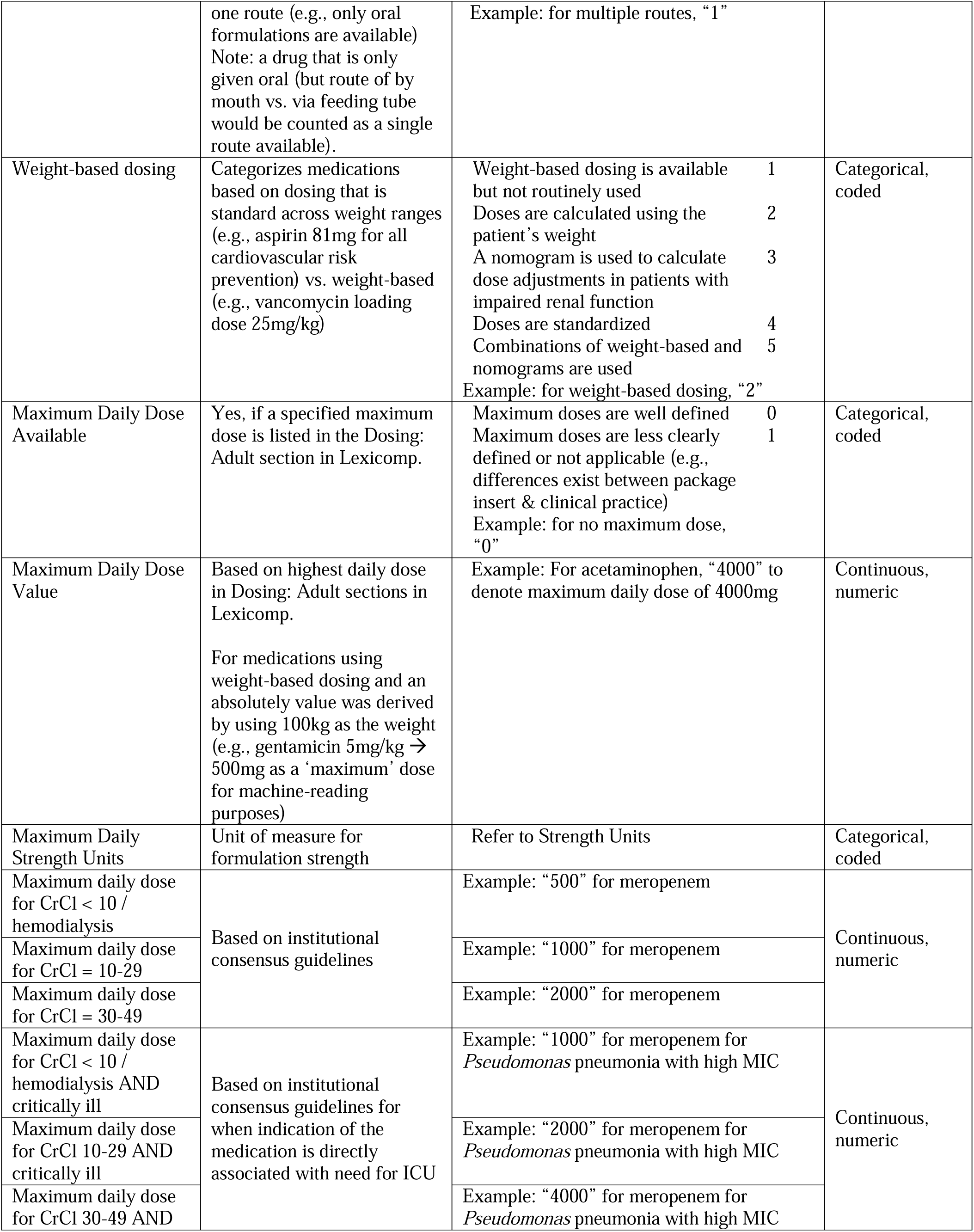
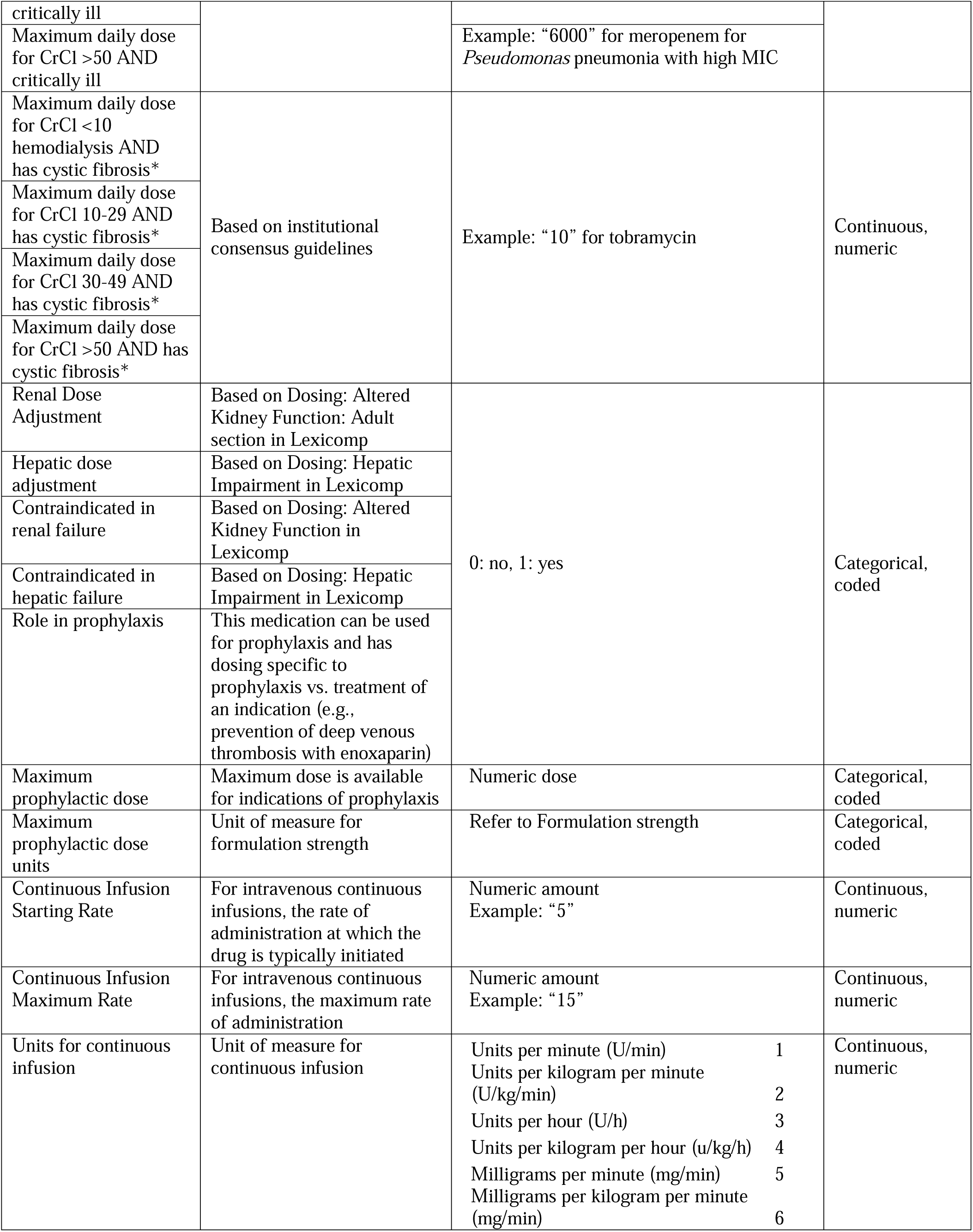
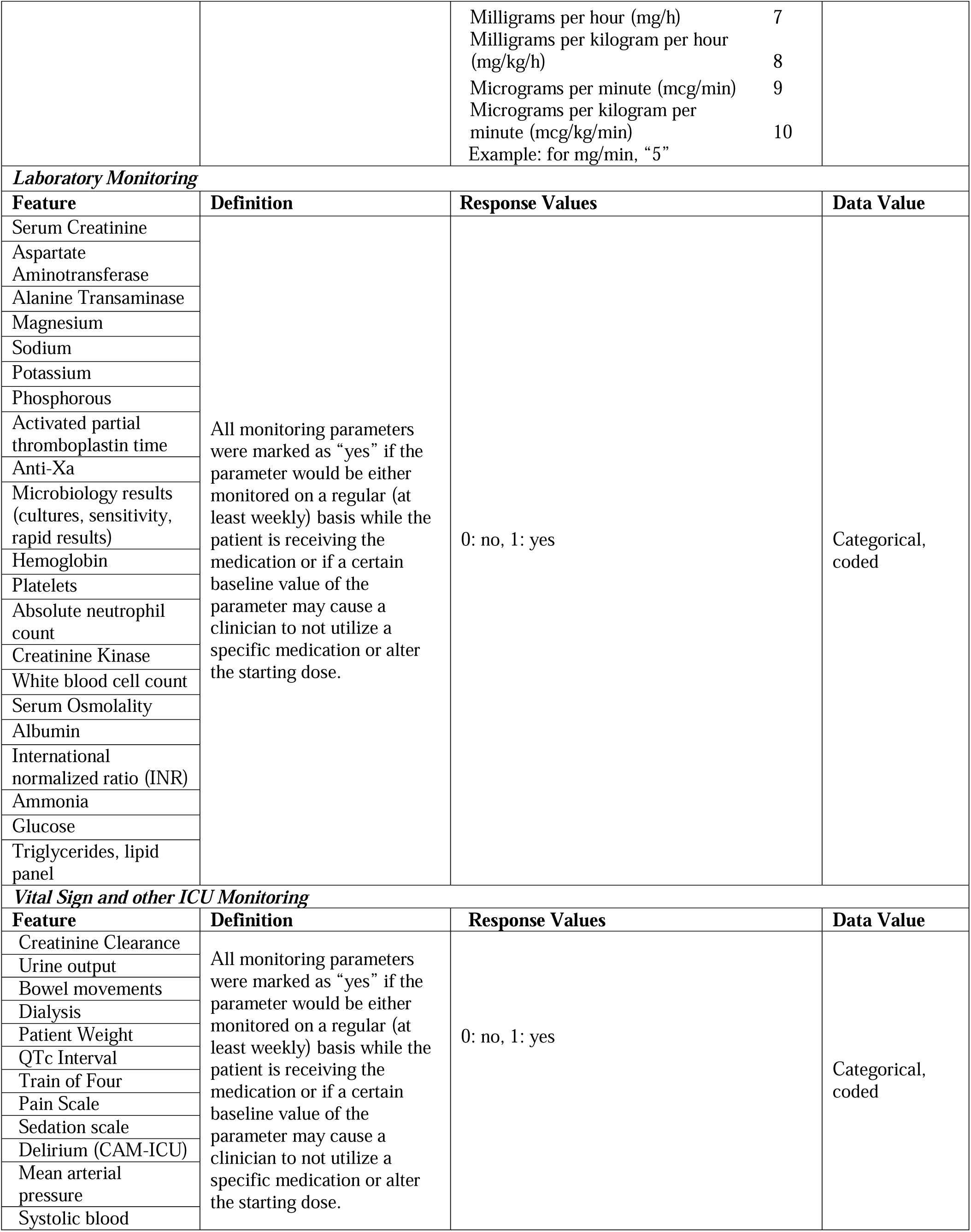
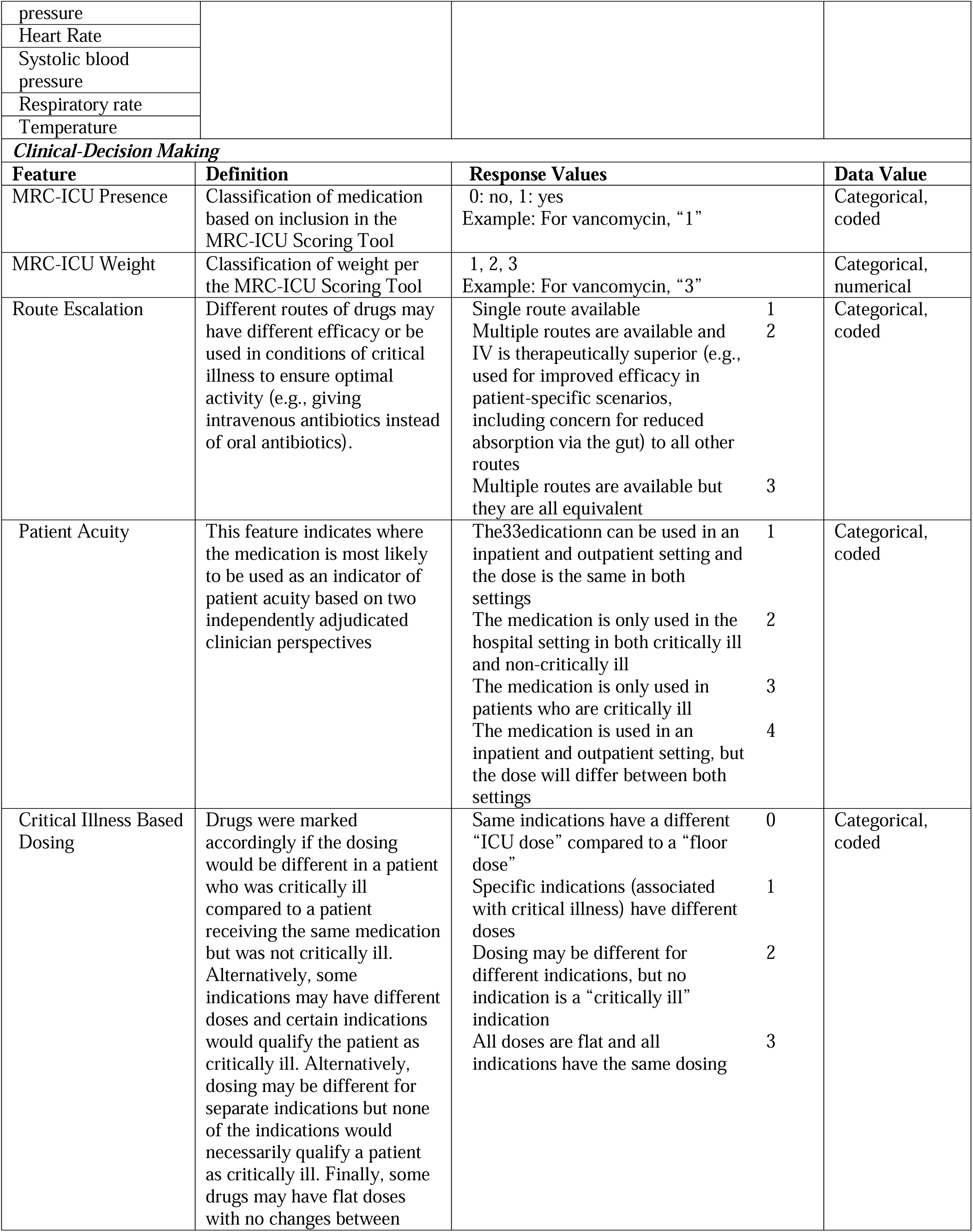
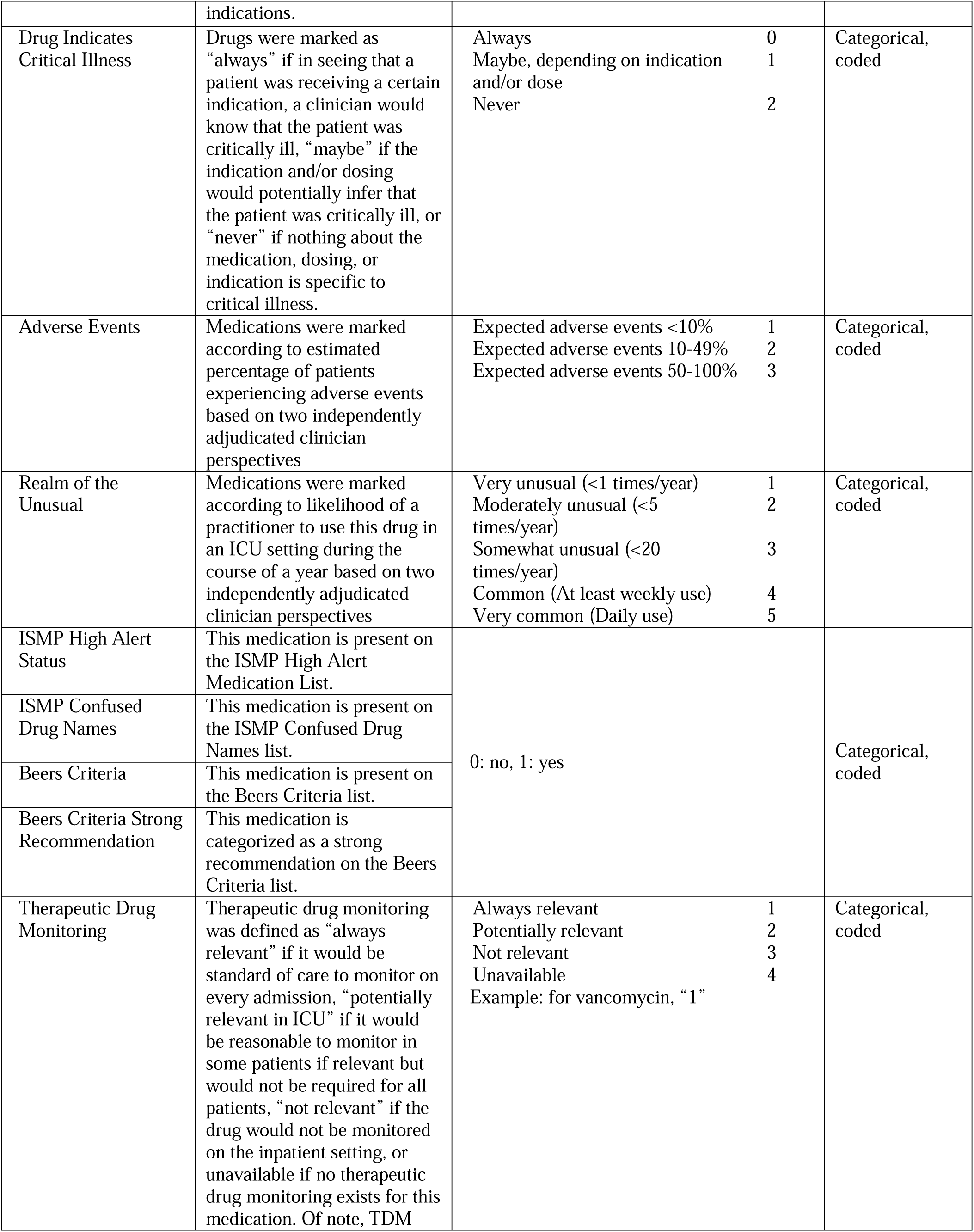
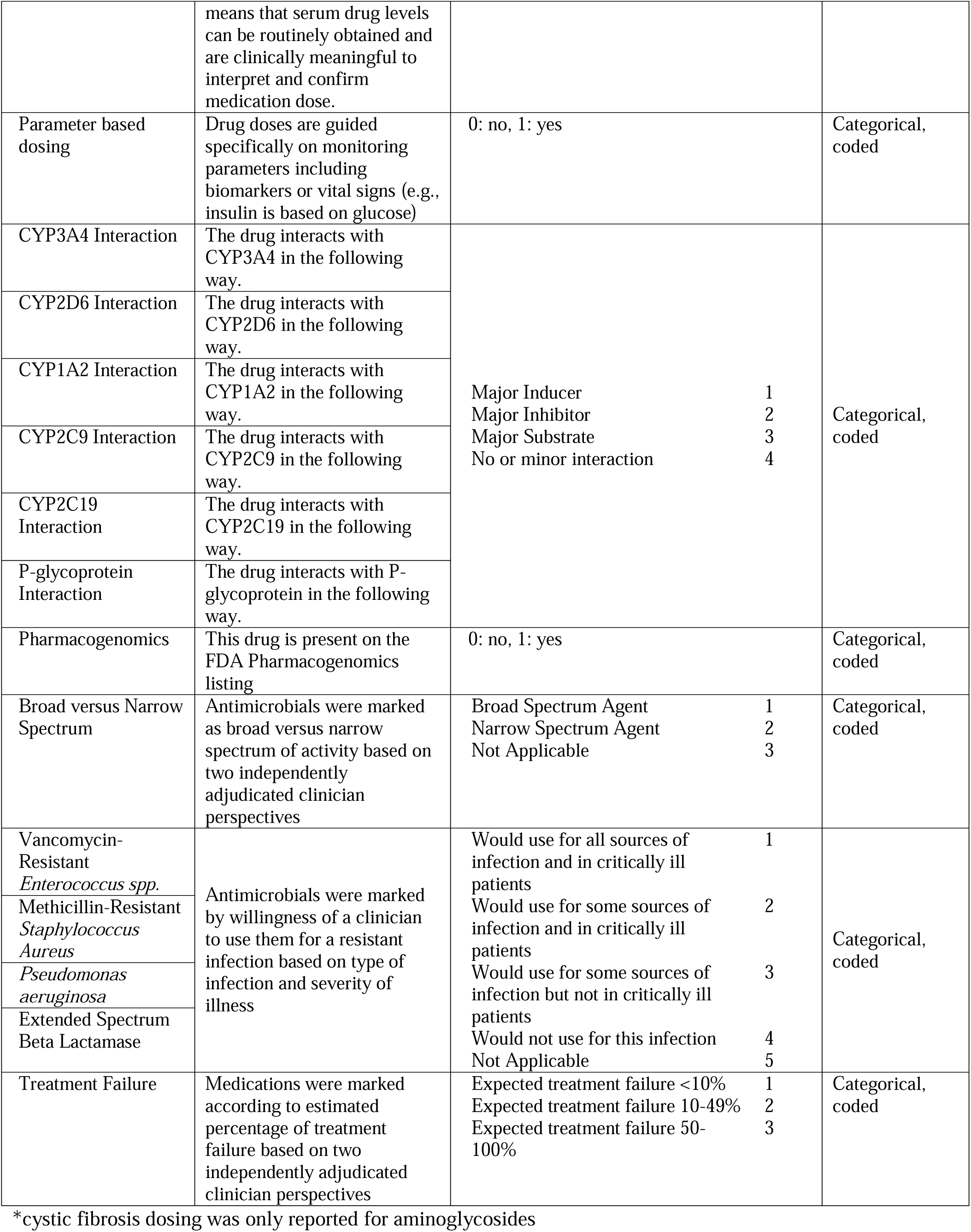
Common Data Model for ICU Medications.

## Contributorship Statement

AS and RK contributed to overall study conception design and interpretation of the work. AS, KK, DM, JD, SS, BM, MB, LC, and SR participated in library design, building, and coding, as relates to substantial contributions to the acquisition and analysis of the work. All authors participated in drafting and critically ill review the work for intellectual content and provided final approval in addition to agreement to be accountable for all aspects of the work.

## Data Availability

The data underlying this article are available in GitHub, under ICURx at: https://www.icurxforai.com/.

## Acknowledgements

Data acquisition were supported by NC TraCS, funded by Grant Number UL1TR002489 from the National Center for Advancing Translations Sciences at the National Institutes of Health, and Data Analytics at the University of North Carolina Medical Center Department of Pharmacy.

American Society of Health-System Pharmacists Innovations in Technology Grant supported this work.

Coding for the common data model was completed with the help of Merrie Barnett, Binh Bui, Alexander Durant, Amber Fraley, Liana Ha, Brittny Nutt, Kara Phillips, and Ciana Wallace.

